# Does diversity beget diversity? A scientometric analysis of over 150,000 studies and 49,000 authors published in high-impact medical journals between 2007 and 2022

**DOI:** 10.1101/2024.03.21.24304695

**Authors:** Marie-Laure Charpignon, João Matos, Luis Nakayama, Jack Gallifant, Pia Gabrielle I. Alfonso, Marisa Cobanaj, Amelia Fiske, Alexander J. Gates, Frances Dominique V. Ho, Urvish Jain, Mohammad Kashkooli, Liam G. McCoy, Jonathan Shaffer, Naira Link Woite, Leo Anthony Celi

**Author notes:** **Code and data availability:** The scripts and datasets underlying this study can be found on our GitHub repository: https://github.com/joamats/mit-scientometrics.

## Abstract

**Background:** Health research that significantly impacts global clinical practice and policy is often published in high-impact factor (IF) medical journals. These outlets play a pivotal role in the worldwide dissemination of novel medical knowledge. However, researchers identifying as women and those affiliated with institutions in low- and middle-income countries (LMIC) have been largely underrepresented in high-IF journals across multiple fields of medicine. To evaluate disparities in gender and geographical representation among authors who have published in any of five top general medical journals, we conducted scientometric analyses using a large-scale dataset extracted from the *New England Journal of Medicine (NEJM)*, *Journal of the American Medical Association (JAMA)*, *The British Medical Journal (BMJ)*, *The Lancet*, and *Nature Medicine*.

**Methods:** Author metadata from all articles published in the selected journals between 2007 and 2022 were collected using the DimensionsAI platform. The Genderize.io API was then utilized to infer each author’s likely gender based on their extracted first name. The World Bank country classification was used to map countries associated with researcher affiliations to the LMIC or the high-income country (HIC) category. We characterized the overall gender and country income category representation across the medical journals. In addition, we computed article-level diversity metrics and contrasted their distributions across the journals.

**Findings:** We studied 151,536 authors across 49,764 articles published in five top medical journals, over a long period spanning 15 years. On average, approximately one-third (33.1%) of the authors of a given paper were inferred to be women; this result was consistent across the journals we studied. Further, 86.6% of the teams were exclusively composed of HIC authors; in contrast, only 3.9% were exclusively composed of LMIC authors. The probability of serving as the first or last author was significantly higher if the author was inferred to be a man (18.1% vs 16.8%, P < .01) or was affiliated with an institution in a HIC (16.9% vs 15.5%, P < .01). Our primary finding reveals that having a diverse team promotes further diversity, within the same dimension (i.e., gender or geography) and across dimensions. Notably, papers with at least one woman among the authors were more likely to also involve at least two LMIC authors (11.7% versus 10.4% in baseline, P < .001; based on inferred gender); conversely, papers with at least one LMIC author were more likely to also involve at least two women (49.4% versus 37.6%, P < .001; based on inferred gender).

**Conclusion:** We provide a scientometric framework to assess authorship diversity. Our research suggests that the inclusiveness of high-impact medical journals is limited in terms of both gender and geography. We advocate for medical journals to adopt policies and practices that promote greater diversity and collaborative research. In addition, our findings offer a first step towards understanding the composition of teams conducting medical research globally and an opportunity for individual authors to reflect on their own collaborative research practices and possibilities to cultivate more diverse partnerships in their work.

## Introduction

Biomedical and public health research that significantly impacts clinical practice and health policy is often contained in high-impact factor (IF) peer-reviewed medical journals^1^. Indeed, several studies have shown that randomized controlled trials (RCT), systematic reviews, and case reports published in high-IF journals, which tend to be highly cited and of superior methodological quality, heavily influence subsequent research, health policy decisions, and implementation at the bedside^2–4^ .

Equity and inclusion are imperative to ensure both the universal and local applicability of published research, while mitigating the risk of adverse safety events among patient minority groups, who are often underrepresented in clinical studies^5^. Research published in high-IF journals should represent the global diversity of health knowledge, policy, and practice, while accounting for the various political, sociocultural, and geographical factors that influence local contexts and may affect the deployment of clinical guidelines. To this end, diversity among authors of research publications should be encouraged to democratize knowledge^6^, to account for underlying social determinants of health in policy recommendations^7^, and to ensure that the research considers the practical needs of underserved populations. Indeed, prior work has shown that collaborative, gender-diverse teams produce higher impact and more innovative scientific research^8^. Similarly, cultural diversity in research teams has positively influenced the quality and quantity of research being produced^9,10^.

Despite the documented benefits of diversity, disparities persist in terms of gender, geographical, and cultural representation of authors. Indeed, women are consistently underrepresented in the fields of science, technology, engineering, and mathematics (STEM)^11^. The same is true in fields other than STEM, including medicine^12–15^ – the focus of our study. Further, across fields, mixed-gender author teams remain significantly underrepresented compared to teams composed only by men^8^. Beyond gender diversity, representation from authors of the Global South is also lacking^16^. In our study, geographical diversity is a shorthand; we use it to quantify the representation of authors affiliated with research institutions located in LMICs vs HICs. This simplified categorization (i.e., LMIC vs HIC status) is used as a proxy to characterize the extent of differences in cultural and socio-economic factors and in access to resources among authors and their institutions, based on their affiliated country. Although LMICs account for 85% of the world’s population and 92% of the global disease burden^17^, articles published in high-IF journals are still predominantly written by authors affiliated with institutions located in HICs. This reality spans multiple fields of medicine, ranging from psychiatry^18^ to surgery^19^ to public health^20^. This discrepancy can be explained by the fact that HICs generally dedicate a higher proportion of their national budget to education^21,22^ as well as to research and development^23,24^ than LMICs; they also have a higher concentration of medical institutions and academic clinical research centers that build a strong(er) health research capacity^25^. To better characterize the current landscape of author diversity in medicine, we set out to assess the diversity of individual investigators and of their scientific publications, in terms of gender identities and geographical areas represented by authors.

We use a scientometric approach to analyze author representation and collaboration patterns in five high-impact and frequently cited general medicine journals. Scientometrics is the quantitative study of science and scholarly research to provide objective data that reflect the impact and relevance of scientific work^26^. The results and interpretation of retrospective scientometric analysis help evaluate the quality of research, influence scientific policies as well as administrative and funding decisions, and identify emerging trends in research topics and methods^27^. Specifically, bibliometric network analysis leverages computational and statistical tools to understand the relationships among co-authors^28^.

We focus on two dimensions: the author’s likely gender and the countries where their institutions are located. Prior studies investigating disparities in author representation have been primarily descriptive, and either journal-specific (i.e., surveying articles from only a single journal or a family of journals)^29–32^, article type-specific (e.g., limiting articles examined to commentaries)^33^, specialty-specific (i.e., surveying articles published in journals of only a specific field)^1,18–20,34,35^, or gap-specific (i.e., focusing on only gender or geographical disparities)^16,36,37^, limiting their conclusions to related contexts. Moreover, prior works have relied primarily on small datasets. For instance, in their observational study of trends in female authors among high-impact medical journals, Filardo et al.^37^ relied on fewer than 4000 articles. Similarly, the analysis of HIC overrepresentation in highly-ranked public health journals by Plancikova et al. was based on fewer than 400 articles^20^. Furthermore, to our knowledge, there is scant literature examining whether – and how – diversity among individual authors affects the extent of homophily in research collaboration networks. In contrast with previous studies, ours takes into account a larger number of journals, is not specialty-or article type-specific, and considers different dimensions of diversity. Uniquely, we quantify how an author’s gender and the income category of the countries where their institutions are located affect authorship position (e.g., being the first or last author). We also analyze how better representation (i.e., the presence of at least one researcher from an underrepresented group, especially in the first or last author position) relates to team composition, within and across dimensions of gender and geographical diversity. We hope that our scientometric analysis of the researchers featured in high-IF medical journals helps promote a more equitable medical knowledge production system.

## Methods

**Figure 1** summarizes the methodological pipeline of this study, which we describe in detail in this section. The scripts and datasets underlying our work can be found on GitHub: https://github.com/joamats/mit-scientometrics.

**Figure 1.**
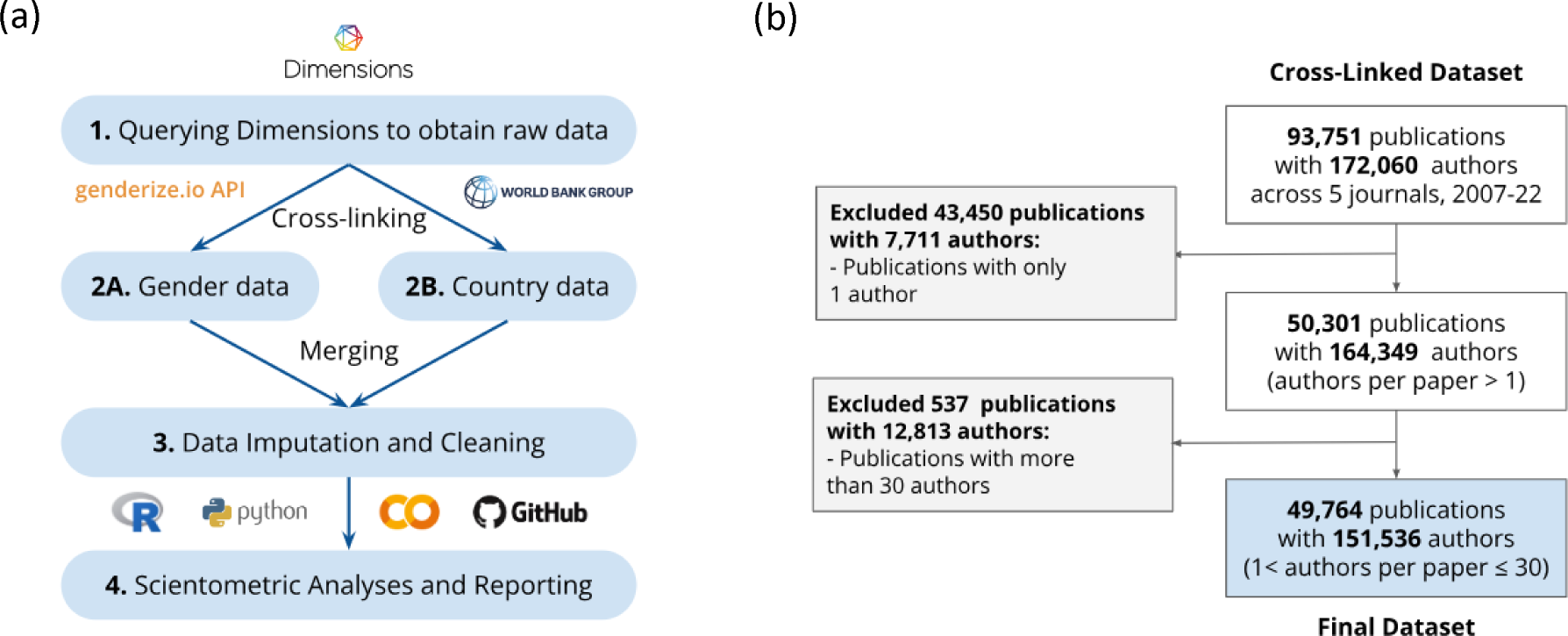
Methodological pipeline to analyze authorship diversity in scientific publications (a) and flow diagram depicting exclusion criteria for publications and authors (b).

### Inclusion criteria and search strategy

This study included metadata about authors of research publications featured in high-IF medical journals. To assess the reach of each medical journal, defined by the average number of citations per research article, we leveraged the Journal Citation Report ranking system from Clarivate Analytics. Specifically, we selected the “General and Internal Medicine” and the “Medicine, Research & Experimental” categories^38,39^. We considered the top five medical journals (see Supplemental Text for more details). We used the DimensionsAI platform, a linked research information dataset, to collect article metadata^40^. We included every article that was indexed by DimensionsAI, had a valid Digital Object Identifier (DOI), and was published in any of the five selected journals between January 2007 and December 2022, irrespective of its type (e.g., observational study, RCT, editorial, systematic review, patient case report). Using a custom query, available in the supplemental text, we retrieved the following information for each article: title, abstract, authors, research affiliations, and countries where research institutions are located.

### Exclusion criteria

To ensure that our analyses focused on collaboration in teams of moderate size, we excluded papers authored by a single author and papers involving more than 30 authors, i.e., 1.07% of the overall dataset. To assess the sensitivity of our results to the choice of this threshold, we conducted two sensitivity analyses, using in turn cut-offs of 40 and 50 authors, corresponding to 0.46% and 0.23% of the overall dataset, respectively.

### Data about authors, research affiliations, and country locations

By cross-linking the publication and author tables available in DimensionsAI, we retrieved the following elements for each author: existing ID, name variants, research affiliations, country locations, research topics, journal publications, co-authors, and years of research activity. Then, each author’s first name was extracted and processed using the Genderize.io Application Programming Interface (API). This API offers access to an underlying database of first names that have been previously labeled as that of a “man” or “woman”. When the number of labeled examples for a given first name is deemed sufficient (i.e., generally 10+ examples), the API returns the probability of an author being a “man” or “woman”; the most likely gender for any given first name can further be determined based on a minimum probability threshold (e.g., 60%). When the number of labeled examples for a given name is deemed insufficient (i.e., generally <10 examples), probabilities cannot be reliably computed; thus, the API returns “unknown” as the most likely gender. Therefore, missing gender data arise from names that are either absent from the underlying database of first names or present but lacking a sufficient number of labeled gender examples. Notably, in the context of our study, gender diversity refers to teams that are not composed exclusively of men or of women. Although a more extensive exploration of gender representation – including that of non-binary and transgender individuals – is warranted, it is beyond the scope of this paper due to data constraints and the challenges associated with gender identity labeling at scale^41^.

Each author of a given paper was also linked to one or multiple countries based on their reported research affiliation(s). For authors who had listed multiple affiliations on a given paper, all corresponding countries were extracted. To analyze the representation of authors affiliated with institutions located in LMICs vs HICs, we considered the latest version of the World Bank Country and Lending Groups dataset, which ranks nations according to their gross national income^42^. We cross-linked the author-country mapping derived from DimensionsAI and the World Bank country-income category mapping, which allowed us to create an additional variable characterizing the proportion of authors affiliated with institutions located in LMICs vs HICs on a given paper.

### Sensitivity analysis to handle missing gender and/or affiliation

We used counterfactual analyses to account for missingness in the inferred gender labels derived from the Genderize API and in the LMIC/HIC-status of authors’ country locations, derived from the *World Bank* dataset. As a baseline approach, we excluded from our study authors with no gender or country income information, assuming that missingness occurs at random. In a first sensitivity analysis, aimed at reflecting a pessimistic scenario, we assumed that all authors with a missing gender (respectively, country income category) label were men (respectively, affiliated with institutions located in HICs). In a second sensitivity analysis, reflecting an optimistic scenario, we assumed the opposite, i.e., all authors with a missing gender (respectively, country income category) label were women (respectively, affiliated with institutions located in LMICs). Authors with multiple affiliations were labeled as being part of institutions located in LMICs only if all their affiliations satisfied this criterion. Importantly, we recognize that LMIC authors may be more likely to have an “unknown” gender label because the databases underlying the Genderize API were built mostly using North American (first name, gender) labeled pairs.

### Statistical analyses

All statistical analyses and figures were generated using Python 3.10 libraries and Google Colaboratory.

First, we sought to answer the following question: “What differentiates top medical journals in terms of the gender and geographical representation of authors?” To this end, we calculated the following quantities for each journal: (1) the proportion of papers with two or more authors that included only researchers affiliated with institutions located in countries of the same income category (either all LMICs or all HICs) and (2) the proportion of papers including a mix of researchers affiliated with institutions located in both LMICs and HICs. Beyond geographical representation, we calculated the proportion of researchers likely to be women, among all authors who published in a given journal during the study period.

Second, we sought to answer the following question: “Does an author’s gender affect their likelihood of being first or last author?”. To this end, we computed the probabilities of being first or last author, conditional on the author’s inferred gender. In addition, we sought to address the following: “Does the income category of the countries where the research institutions of a given author are located affect their likelihood of being first or last author?”. We similarly computed the probabilities of being first or last author, conditional on the author being affiliated with research institutions located in LMICs vs HICs. In total, we tested four null hypotheses: (a, b) an author’s gender does not affect their likelihood of being first (a) or last (b) author; (c, d) the income category of the countries where the research institutions of a given author are located does not affect their likelihood of being first (c) or last (d) author. Notably, in the field of medicine, the first author is often in charge of designing the study, conducting the analysis, writing the first draft of the manuscript, or all of them; the last author is often the principal investigator facilitating access to data or other infrastructural resources, overseeing the conduct of the study, providing guidance about manuscript writing, or all of them.

We used regular bootstrap with resampling (100 iterations) to compute 95% confidence intervals for each proportion and conditional probability. To evaluate the plausibility of the null hypotheses, we used chi-squared tests. We used Bonferroni correction to account for multiple hypothesis testing; null hypotheses were rejected for p-values less than 0.0125, i.e., a familywise error rate of 0.05 by 4, the number of executed tests.

Finally, to answer the question “Does diversity beget diversity?”, we studied the composition of authoring teams in different scenarios. We hypothesized that having a more diverse team – i.e., including a greater mix of authors from over- and under-represented groups – would beget further diversity, both within the same dimension and across dimensions. To test this hypothesis, we split the dataset into six non-exclusive, possibly overlapping groups of publications, based on whether their authoring team met the following conditions: “the first author is likely a woman” (group 1); “the first author is affiliated with an LMIC institution” (group 2); “the last author is likely a woman” (group 3); “the last author is affiliated with an LMIC institution” (group 4); “at least one author is likely a woman” (group 5); “at least one author is affiliated with an LMIC institution” (group 6); and then studied team composition, similarly. Notice that a team can have both their first and last authors be affiliated with an LMIC institution, in which case the corresponding publication would be eligible to enter both groups 3 and 4. Null hypotheses for each of the 6 aforementioned tests were rejected at 0.0083, after applying a Bonferroni correction for a familywise error rate of 0.05.

In the Results section that follows, we report the findings emanating from our main analysis. The results corresponding to the various sensitivity analyses that we implemented can be found in the Supplementary Materials.

## Results

### Overview of the dataset

The 2021 impact factor (IF) of the five selected journals, as defined by Clarivate Analytics, ranged from 87.2 (*Nature Medicine*) to 202.7 (*The Lancet*)^43^. See **Supplemental Table 1** for a summary of the journals’ IF and **Supplemental Figure 1** for their evolution over time.

Our initial query yielded 93,751 publications and 172,060 authors. After applying our exclusion criteria, based on the number of authors of the publications, we obtained 49,764 publications from a total of 151,536 authors, across the five studied journals, from 2007-2022. The exact publication date and title were available for all papers included in the analysis. **Figure 1b** depicts the consort diagram of our study cohort.

### Summary statistics about publications, across journals and years

Together, the BMJ and The Lancet accounted for over half of the articles published during the study period, with a total of 11,481 (23.1%) and 16,400 (33.0%) publications, respectively. The remaining papers were split among JAMA, 9,886 (19.9%), NEJM, 8,303 (16.7%), and Nature Medicine, 3,694 (7.4%). Our study period spans 15 years; however, the number of yearly publications has increased in more recent years, with the 2019-2022 time period alone accounting for 18,542 articles (37.3%). See **Table 1** for details on the distribution of the research output across journals and years.

**Table 1.** Summary statistics about publications.

|  |  | **Overall** |
| --- | --- | --- |
| **N** |  | 49,764 |
| **Journal,**<br>**N (%)** | **JAMA** | 9,886 (19.9) |
|  | **Nature Medicine** | 3,694 (7.4) |
|  | **NEJM** | 8,303 (16.7) |
|  | **The BMJ** | 114,81 (23.1) |
|  | **The Lancet** | 16,400 (33.0) |
| **Years,**<br>**N (%)** | **2007-2010** | 6,590 (13.2) |
|  | **2011-2014** | 10,673 (21.4) |
|  | **2015-2018** | 13,959 (28.1) |
|  | **2019-2022** | 18,542 (37.3) |

### Gender and country income diversity in top medical journals

A descriptive analysis of the authors included in the dataset can be found in **Table 2**. Using the *Genderize.io API*, we identified 51,150 authors (33.8%) as likely to be women and 85,967 authors (56.7%) as likely to be men; the remaining 14,419 authors (9.5%) could not be mapped to either group and were thus subsequently labeled as missing. As for the country income of authors’ affiliations, 132,599 (87.5%) were affiliated with an institution from a HIC and 16,322 (10.8%) with an institution from an LMIC; the remaining 2,615 (1.7%) authors could not be mapped to either category and were thus subsequently labeled as missing. As a result, 5,172 (10.4%) papers had at least one LMIC author. Notably, we applied a strict criterion for authors to be considered as affiliated with an LMIC, i.e., all of their institutions had to be located in an LMIC. In total, 87,980 authors (47.9%) had more than one affiliation; 6,238 authors (7.1%) had all of their affiliations mapping to an LMIC; and 13,301 (15.1%) authors had at least one such affiliation.

**Table 2.** Summary statistics about authors (with no imputation for missing gender and country income category).

|  | Grouped by gender |  |  |  | Grouped by country income |  |  |  |
| --- | --- | --- | --- | --- | --- | --- | --- | --- |
|  | Missing | Male | Female | P-Value | Missing | HIC | LMIC | P-Value |
| **N (%)** | 14,419 (9.5) | 85,967 (56.7) | 51,150 (33.8) |  | 2,615 (1.7) | 132,599 (87.5) | 16,322 (10.8) |  |
| **1+ first-authored paper, N (%)** |  | 13,617 (15.8) | 7,443 (14.6) | <0.001 |  | 20,336 (15.3) | 2,419 (14.8) | 0.086 |
| **1+ last-authored paper, N (%)** |  | 13,829 (16.1) | 7,527 (14.7) | <0.001 |  | 20,477 (15.4) | 2,499 (15.3) | 0.667 |
| **2+ first-authored paper, N (%)** |  | 5,164 (6.0) | 2,213 (4.3) | <0.001 |  | 7,220 (5.4) | 616 (3.8) | <0.001 |
| **2+ last-authored paper, N (%)** |  | 5,071 (5.9) | 2,187 (4.3) | <0.001 |  | 7,112 (5.4) | 604 (3.7) | <0.001 |
| **Published in 2+ journals, N (%)** |  | 16,512 (19.2) | 6,930 (13.5) | <0.001 |  | 22,975 (17.3) | 1,921 (11.8) | <0.001 |
| **Published 2+ papers, N (%)** |  | 29,189 (34.0) | 14,383 (28.1) | <0.001 |  | 42,528 (32.1) | 4,374 (26.8) | <0.001 |

\* LMIC: low- and middle-income country, HIC: high-income country.

Researchers inferred to be men and individuals affiliated with an institution located in an HIC accounted for a larger share of publications than those inferred to be women and those from an LMIC.

Trends in gender and geographical representation remained robust to the imputation strategy used to handle missing gender and missing country information. In **Supplemental Table 2a** (respectively, **Supplemental Table 2b**), we provide the results of a sensitivity analysis where we assumed all authors with a missing gender label were women (respectively, men). Similarly, in **Supplemental Table 3a** (respectively, **Supplemental Table 3b**), we provide the results of a sensitivity analysis where we assumed all authors with a missing LMIC/HIC-status were affiliated with an institution located in an LMIC (respectively, HIC). Note that we used these optimistic and pessimistic imputation strategies to derive bounds and avoid assuming that gender labels were missing at random.

In **Supplemental Figure 2a**, we provide the distribution of authors by continent. We show that most authors were affiliated with institutions located in Europe (40.5%) and North America (39.4%), followed by Asia (10.8%), Oceania (3.7%), Africa (2.7%), and South America (1.5%). The distribution by country is provided in **Supplemental Figure 2b**, limited to the top 20 countries out of 196 distinct countries represented in the overall dataset. A sizable fraction of authors was affiliated with at least one institution located in the United States (US; 34.9%) or Great Britain (GB; 7.1%).

### Gender and geographical diversity at the team level

Team diversity was assessed in terms of the most likely gender of the authors and of the income level of the country where their institution(s) were located (**Supplemental Figure 2**).

**Figure 2a** depicts gender diversity and compares the performance of the five journals. We set 50% as the gender parity threshold. Gender parity was not achieved by any journal, even when assuming that all authors with missing gender labels were women. Overall, 33.1% of the authors in each team were likely women. Gender representation was the lowest for NEJM publications (31.0%) and the highest for BMJ publications (34.9%).

**Figure 2.**
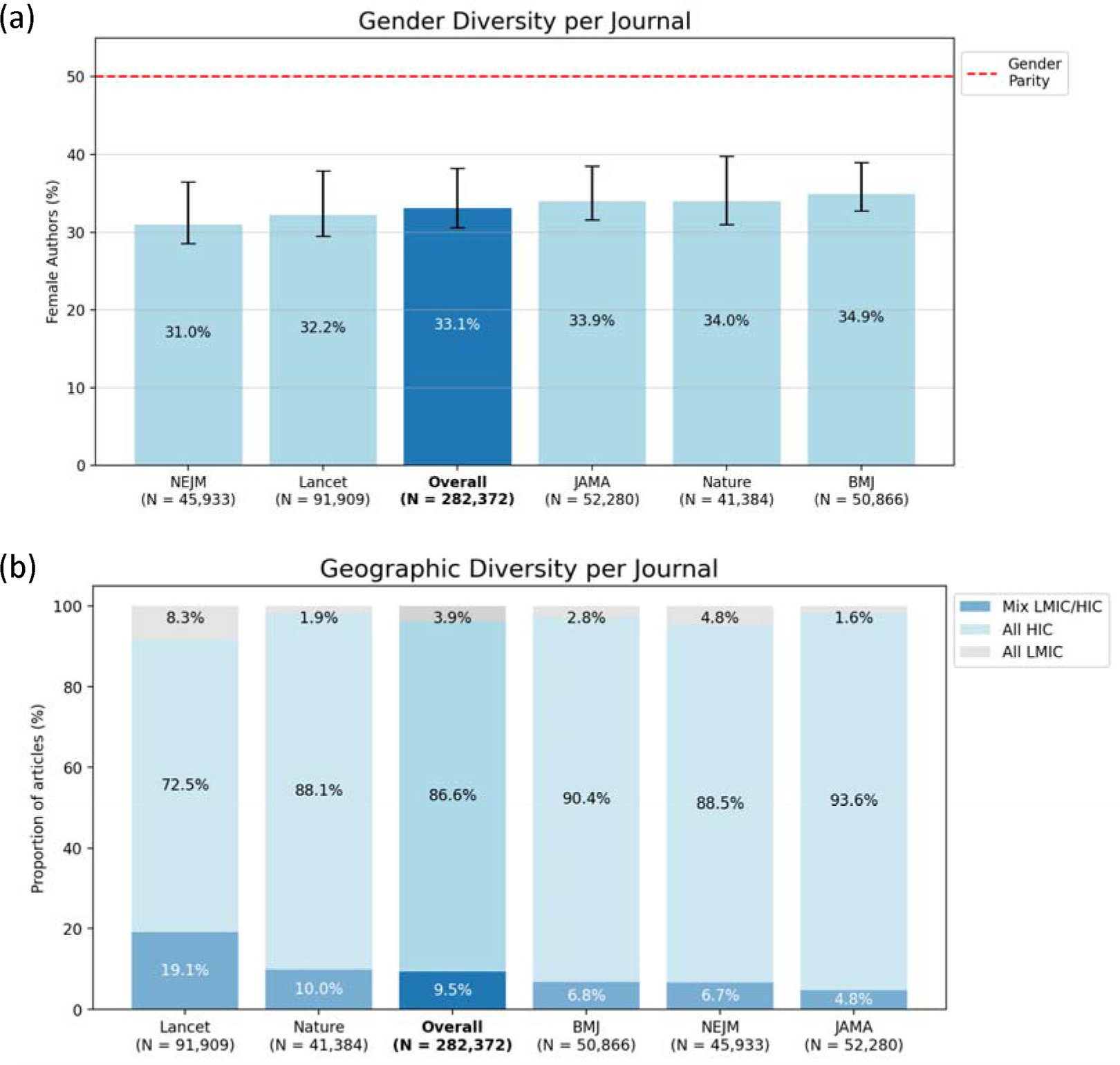
Gender (a) and geographical (b) diversity, in terms of team composition, stratified by journal. (a) Bars represent the scenario in which authors with missing gender information are removed from the dataset, while error ranges represent the optimistic and pessimistic imputation strategies described in the Methods section. (b) All numbers refer to the scenario in which authors with missing information are excluded; pessimistic and optimistic scenarios can be found in the **Supplemental Figures 3a and 3b**.

In **Supplemental Tables 4a, 4b, and 4c**, we provide sensitivity analyses to the maximum number of authors per article included in the study (30, 40, 50). Our findings remained consistent, both overall and by journal.

Next, we characterized team diversity in terms of country income categories represented by the authors (**Figure 2b**). Overall, we found that most teams were homophilous: 86.9% (respectively, 3.9%) were composed exclusively of authors affiliated with institutions located in HICs (respectively, LMICs). Only 9.5% of teams included both HIC and LMIC authors. *JAMA*, *NEJM*, and *The BMJ* had lower team-level geographical representation (4.8%, 6.7%, and 6.8% of teams had both LMIC and HIC authors., respectively), while *Nature Medicine* and *The Lancet* had higher representation (10.0% and 19.1% of the teams had both LMIC and HIC authors, respectively).

Our sensitivity analyses, reflecting optimistic and pessimistic scenarios for the imputation of missing information on country income, can be found in **Supplemental Figures 3a and 3b**.

**Figure 3.**
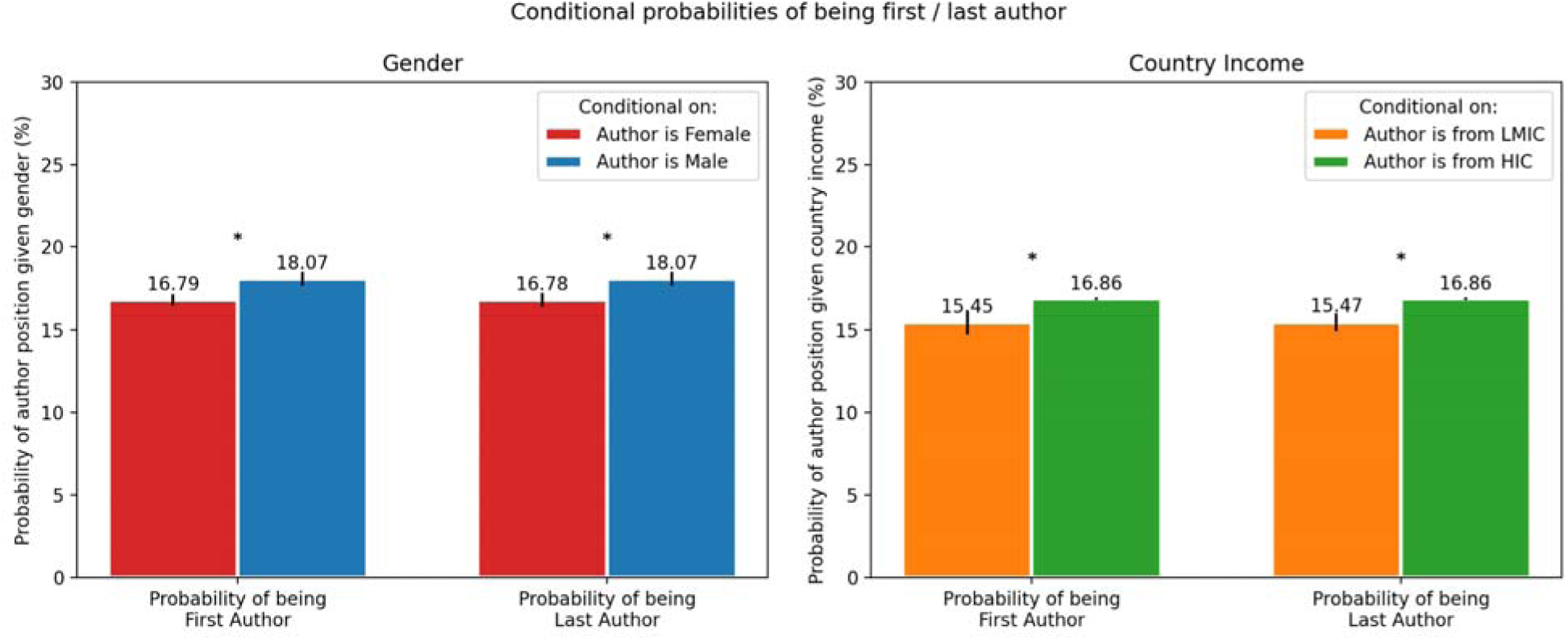
Conditional probabilities of being first and last author given author’s gender and the LMIC-status of their affiliation. The asterisk (*) indicates a statistically significant difference between two groups (e.g., author *is fro m LMIC* versus *is from HIC*), with a p-value less than 0.05/4 = 0.0125, as determined by a Chi-squared test, after applying a Bonferroni correction for a familywise error rate of 0.05 and 4 tests. The error bars represent a 95% confidence interval, computed via 100-iteration bootstrap.

### Authorship position

Regarding authorship position, our unadjusted analyses show that being a man or being affiliated with an institution from an HIC increased the probability of being first or last author. We report these findings in **Figure 3**, using conditional probabilities.

Authors inferred to be women had a probability of 16.79% (95% CI, 16.43 – 17.15%) of being first author and 16.78% (95% CI, 16.37 – 17.21%) of being last author. Authors inferred to be men were more likely to take such roles: 18.07% (95% CI, 17.87 – 18.27%) and 18.07% (95% CI, 17.86 – 18.28%), respectively. The estimated difference among likely gender groups was statistically significant (P < 0.0125).

Similarly, researchers affiliated with an institution located in an LMIC had a 15.45% (95% CI, 14.75 – 16.15%) probability of taking a first author role and a 15.47% (95% CI, 14.96 – 15.98%) probability of taking a last author role. In contrast, researchers whose institution was based in a HIC had a 16.86% (95% CI, 16.77 – 16.95%) and 16.86% (95% CI, 16.77 – 16.95%) probability of taking such roles, respectively. The estimated difference among income country groups was statistically significant (P < 0.0125).

### Diversity begets diversity

We show that, when a team had at least one author who was likely a woman or had at least one LMIC researcher, then the other authors were also more likely to be from underrepresented groups, so the team’s overall composition was more diverse. In **Figure 4**, we provide a selected set of conditional probabilities supporting this finding, i.e., diversity begets diversity. The left heatmap refers to gender diversity, while the right heatmap relates to country income diversity. The first column of each heatmap, which represents baseline probabilities with no conditioning of any kind, can be used as a reference. In particular, its values can be compared with those in the second and third columns, which represent conditional probabilities. Moving from left to right, the denominator decreases, which is the reason why different cells with the same numerator value may have different probabilities. Baseline probabilities were derived using the entirety of publications. Conditional probabilities were calculated based on publications with at least one author likely to be a woman or with at least one LMIC author. Importantly, the relationships encoded in the two heatmaps are associative in nature rather than causal.

**Figure 4.**
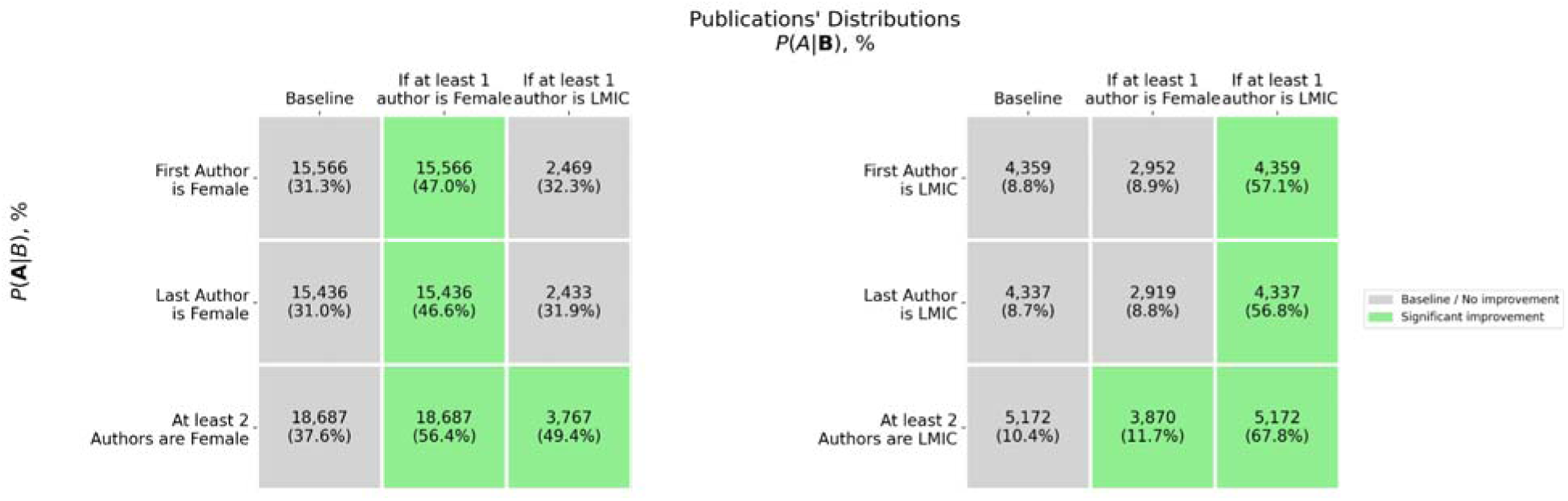
Selected conditional probabilities to assess the impact of team composition in its diversity. Green cells indicate a statistically significant difference between groups (e.g., *there is at least one female author in the team* versus *there is no female author*), with a p-value less than 0.05/6 = 0.0083, as determined by a Chi-squared test, after applying a Bonferroni correction for a familywise error rate of 0.05 and 6 tests.

First, we found that publications with at least one author who was likely a woman had more diverse teams overall. Specifically, the presence of at least one author who was likely a woman increased the probability that the first author was likely a woman (47.0% versus 31.3% in baseline, P < 0.001); that the last author was likely a woman (46.6% versus 31.0% in baseline, P < 0.001); that there were at least two authors who were also likely women (56.4% versus 37.6% in baseline, P < 0.001); and that there were at least two LMIC authors (11.7% versus 10.4% in baseline, P <0.001) in the team. However, the presence of at least one author who was likely a woman did not significantly improve the probability of having a first or last author affiliated with an institution located in an LMIC.

Second, we found that publications with at least one LMIC author also had more diverse teams overall. Specifically, the presence of at least one LMIC author increased the probability that the first author was affiliated with an institution located in an LMIC (57.1% versus 8.8% in baseline, P < 0.001); that the last author was from an LMIC (56.8% versus 8.7%, in baseline, P < 0.001); that there were at least two LMIC authors (67.8% versus 10.4%, in baseline, P < 0.001); and that there were at least two authors who were likely women (49.4% versus 37.6%, P < 0.001). However, the presence of at least one LMIC author on the team did not significantly improve the probability of having an author who was likely a woman as first or last author.

In **Supplemental Table 5**, we present the complete set of results for this section. For example, we explored additional conditions on the composition of the authoring team, such as having as the first or last author a researcher who is likely a woman or who is affiliated with an institution located in a LMIC. These investigations complement the results presented in the above paragraphs, which do not consider authorship role/position. Given the large number of tests we ran, we set the significance threshold to a lower value to account for multiple hypothesis testing and thus minimize the number of false positive findings; nevertheless, the findings about gender and country income diversity remained the same.

Overall, we show that having at least one member from an underrepresented group promoted further diversity in the authoring teams we studied, in terms of likely gender, country income category, or both. Notably, the most interesting finding emerged from our cross-dimension analyses, which revealed that having LMIC representation in the team simultaneously promoted the representation of authors who were likely women, and vice-versa.

## Discussion

Our large-scale analysis of over 49,000 publications and 150,000 authors shows that the five high-impact medical journals included in this study lack diverse gender and geographical representation. Only a fraction of research works featured authors who were likely women (33.1%) or who were affiliated with institutions located in LMICs (including teams exclusively composed of LMIC authors, as well as teams that featured at least one LMIC author, totaling to 13.4%). This finding corroborates prior literature^20^ and underscores the concerning persistence of the underrepresentation of marginalized voices^11–14,16,20,44^. Notably, journals with a broader or global health focus, such as the BMJ and the Lancet, featured more diverse teams than the other three journals, suggesting that the inclusion of authors from underrepresented groups in reputable medical and health sciences venues was achievable. This research offers a foundational framework for scientometrics, spotlighting the dire need for greater diversity, more balanced representation of genders, and stronger collaboration among countries of different income levels. Through the open-access release of a dataset of unprecedented size, we aim to prompt academic journals, editorial boards, and research institutions to take a closer look at the voices represented in the studies they review and/or publish and to proactively revisit their inclusion strategies.

### Compounding diversity

A key takeaway from our study is the potential synergy between gender and geographical diversity^45^. We found that publications whose first or last author was likely a woman also had a higher proportion of LMIC contributors. While our current study design does not allow causal interpretations, the findings suggest that fostering gender diversity in academic collaborations could act as a catalyst for greater geographical representation. We hypothesize that the confluence of diverse gender perspectives within a team may emphasize the need for the authors to reach out to local experts, leading in turn to more global collaborations in medicine and health sciences. Future studies should further investigate this synergy and consider other forms of diversity like race, ethnicity, and sexual orientation. Ultimately, championing diversity could accelerate the democratization of research and thus broaden the scientific knowledge base.

### Limitations

Despite the robustness of our study findings to the approaches we used to handle missing gender and country income information, a few limitations must be acknowledged. A crucial limitation is the binary treatment of gender, which is necessitated by the pre-trained algorithm from Genderize.io that we used. This algorithmic simplification ignores the fact that gender is multifaceted; any assumption based on names commonly seen in certain contexts is probabilistic in nature and can misrepresent non-binary gender identities. The absence of any self-identified gender data, which would have been challenging to collect due to the large sample size, prevents us from evaluating the extent of this limitation. Because the current methodology risks oversimplifying the diverse gender spectrum, we have been careful in using the language of authors who are “*likely”* or “inferred” to be a particular gender throughout the article.

Furthermore, our results on increased diversity in teams including an author from an underrepresented group are associative in nature and should not be interpreted causally. Unmeasured confounders such as the seniority of authors and historical hiring practices in certain institutions may partially alter our results. Thus, future efforts should focus on a more precise characterization of researchers and on causal and counterfactual analyses to improve our understanding of the underlying team dynamics, overall and in the presence of marginalized voices.

Finally, our study focuses on team diversity in the field of medicine, limiting our ability to extrapolate to other disciplines. For example, whether our results generalize to the biomedical sciences, epidemiology, and public health remains to be determined. Future work should investigate the extent of similarities and differences in author representation among such adjacent fields.

### Missed opportunities and the value of collective expertise in academic research

The implications of our study findings are multifaceted. While our finding about the synergy between gender and geographical diversity is encouraging, improvements in representation among authoring teams necessitate structural reforms and investments. These efforts are essential to sustainably support both the training of a growing pool of medical and health researchers in LMICs^46,47^ and the professional development of women in LMICs and HICs alike.

Indeed, the lack of diversity and inclusion in academic authorship signifies a missed opportunity to bring together varied perspectives and expertise. In the long run, this lack can lead to the production of a narrowed body of medical knowledge. If medical and health research is not inclusive, its practical applicability to diverse patient populations can be limited, impacting clinical and policy decisions as well as patient care globally. Moreover, the risk of replicating pernicious forms of social bias within research design and practice may be higher when none of the authors can speak to the experience of underrepresented groups or has worked within medical systems that differ from those in HICs. Societally, the absence of gender parity and the limited involvement of researchers affiliated with institutions located in LMICs can influence the topics of global health research, to the detriment of patient populations living in LMICs. Furthermore, diversity in research authorship is a matter of science diplomacy: not only does it enrich the research landscape but it also advances scientific and technological innovation, e.g., via international partnerships. That is why significant social and economic investments, nationally and internationally, are essential to foster opportunities that would ultimately yield greater academic diversity. Key policies should focus on promoting equal access to education programs^48^ and fair treatment within academia^49^, aiming for further advances in competitiveness^50^, inventiveness^8^, and productivity^43,51^. In this pursuit, special consideration should be given to involving intersectional identities and to sponsoring continued education, locally or in partner countries. For instance, women affiliated with institutions located in LMICs may strongly benefit from educational and research training programs conducted in their home institutions or elsewhere. Across country income categories, professional development should be encouraged for doctoral students and established researchers alike, with an emphasis on knowledge transmission across generations. Tools such as massive open online courses^52^ and shadowing opportunities could be leveraged to that end. Given the uneven distribution of research and training resources between HICs and LMICs, we contend that researchers affiliated with well-equipped institutions should spur the effort of cultivating team diversity through internal and external grants.

## Conclusion

Our work highlights that diversity in authorship, including through research leaders from underrepresented groups, can beget further team diversity, and that initiatives encouraging greater gender and geographical representation among authors may be synergistic. We hope this study serves as a step towards enhancing gender parity and LMIC representation in medical and health research collaborations, thereby enriching the breadth and depth of global medical knowledge.

## Data Availability

The scripts and datasets underlying this study can be found on our GitHub repository: https://github.com/joamats/mit-scientometrics

## References

1. Bojanic T, Tan AC. International representation of authors, editors and research in neurology journals. BMC Med Res Methodol. 2021;21(1):57. doi:10.1186/s12874-021-01250-9

2. Albrecht J, Meves A, Bigby M. Case reports and case series from Lancet had significant impact on medical literature. J Clin Epidemiol. 2005;58(12):1227–1232. doi:10.1016/j.jclinepi.2005.04.003

3. Fleming PS, Koletsi D, Seehra J, Pandis N. Systematic reviews published in higher impact clinical journals were of higher quality. J Clin Epidemiol. 2014;67(7):754–759. doi:10.1016/j.jclinepi.2014.01.002

4. Catalá-López F, Aleixandre-Benavent R, Caulley L, et al. Global mapping of randomised trials related articles published in high-impact-factor medical journals: a cross-sectional analysis. Trials. 2020;21(1):34. doi:10.1186/s13063-019-3944-9

5. Chauhan A, Walton M, Manias E, et al. The safety of health care for ethnic minority patients: a systematic review. Int J Equity Health. 2020;19(1):118. doi:10.1186/s12939-020-01223-2

6. Masuda JR, Creighton G, Nixon S, Frankish J. Building Capacity for Community-Based Participatory Research for Health Disparities in Canada: The Case of “Partnerships in Community Health Research.” Health Promot Pract. 2011;12(2):280–292. doi:10.1177/1524839909355520

7. El Ansari W. Collaborative research partnerships with disadvantaged communities: challenges and potential solutions. Public Health. 2005;119(9):758–770. doi:10.1016/j.puhe.2005.01.014

8. Yang Y, Tian TY, Woodruff TK, Jones BF, Uzzi B. Gender-diverse teams produce more novel and higher-impact scientific ideas. Proc Natl Acad Sci. 2022;119(36):e2200841119. doi:10.1073/pnas.2200841119

9. Stahl GK, Maznevski ML. Unraveling the effects of cultural diversity in teams: A retrospective of research on multicultural work groups and an agenda for future research. J Int Bus Stud. 2021;52(1):4–22. doi:10.1057/s41267-020-00389-9

10. Hattery AJ, Smith E, Magnuson S, et al. Diversity, Equity, and Inclusion in Research Teams: The Good, The Bad, and The Ugly. Race Justice. 2022;12(3):505–530. doi:10.1177/21533687221087373

11. UNESCO Institute for Statistics. Women in Science. Published online June 2019. Accessed December 1, 2023. https://uis.unesco.org/sites/default/files/documents/fs55-women-in-science-2019-en.pdf

12. Sela N, Anderson BL, Moulton AM, Hoffman AL. Gender Differences in Authorship Among Transplant Physicians: Are We Bridging the Gap? J Surg Res. 2021;259:271–275. doi:10.1016/j.jss.2020.09.037

13. Tran TB, Wong P, Raoof M, Melstrom K, Fong Y, Melstrom LG. The evolving gender distribution in authorship over time in American surgery. Am J Surg. 2022;224(5):1217–1221. doi:10.1016/j.amjsurg.2022.05.029

14. Abdalla M, Abdalla M, Abdalla S, Saad M, Jones DS, Podolsky SH. The Under-representation and Stagnation of Female, Black, and Hispanic Authorship in the Journal of the American Medical Association and the New England Journal of Medicine. J Racial Ethn Health Disparities. 2023;10(2):920–929. doi:10.1007/s40615-022-01280-z

15. Huang J, Gates AJ, Sinatra R, Barabási AL. Historical comparison of gender inequality in scientific careers across countries and disciplines. Proc Natl Acad Sci. 2020;117(9):4609–4616. doi:10.1073/pnas.1914221117

16. Ghani M, Hurrell R, Verceles AC, McCurdy MT, Papali A. Geographic, Subject, and Authorship Trends among LMIC-based Scientific Publications in High-impact Global Health and General Medicine Journals: A 30-Month Bibliometric Analysis: J Epidemiol Glob Health. 2020;11(1):92. doi:10.2991/jegh.k.200325.001

17. Global Forum for Health Research. The 10/90 Report on Health Research 2003-2004. www.globalforumhealth.org

18. El Khoury J, Kanj R, Adam L, et al. The contribution of authors from low- and middle-income countries to top-tier mental health journals. Eur Sci Ed. 2021;47:e72187. doi:10.3897/ese.2021.e72187

19. Mahawar KK, Kumar G, Malviya A. Who Publishes in Leading General Surgical Journals? The Divide Between the Developed and Developing Worlds. Asian J Surg. 2006;29(3):140–144. doi:10.1016/S1015-9584(09)60073-7

20. Plancikova D, Duric P, O’May F. High-income countries remain overrepresented in highly ranked public health journals: a descriptive analysis of research settings and authorship affiliations. Crit Public Health. 2021;31(4):487–493. doi:10.1080/09581596.2020.1722313

21. The World Bank. The Adequacy Of Public Expenditure On Education And The Needs Post-Covid-19.; 2023. https://thedocs.worldbank.org/en/doc/9b9ecb979e36e80ed50b1f110565f06b-0200022023/original/Adequacy-Paper-Final.pdf

22. The World Bank and UNESCO. Education Finance Watch 2022.; 2022. http://creativecommons.org/licenses/by-sa/3.0/igo/

23. World Intellectual Property Organization. Global Innovation Index 2021: Tracking Innovation through the COVID-19 Crisis.; 2021.

24. World Intellectual Property Organization. The Global Innovation Index 2020.; 2020.

25. Kilmarx PH, Maitin T, Adam T, et al. Increasing Effectiveness and Equity in Strengthening Health Research Capacity Using Data and Metrics: Recent Advances of the ESSENCE Mechanism. Ann Glob Health. 2023;89(1). doi:10.5334/aogh.3948

26. Goldenberg D. Scientometrics and its positive consequences. Rev Bras Cir Plástica RBCP – Braz J Plast Sugery. 2017;32(4):471–471. doi:10.5935/2177-1235.2017RBCP0077

27. Sohn E, Noh KR, Lee B, Kwon OJ. Bibliometric Network Analysis and Visualization of Research and Development Trends in Precision Medicine. In: 2018 IEEE/ACM International Conference on Advances in Social Networks Analysis and Mining (ASONAM). IEEE; 2018:727–730. doi:10.1109/ASONAM.2018.8508350

28. Mejia C, Wu M, Zhang Y, Kajikawa Y. Exploring Topics in Bibliometric Research Through Citation Networks and Semantic Analysis. Front Res Metr Anal. 2021;6:742311. doi:10.3389/frma.2021.742311

29. González-Alvarez J. Author gender in The Lancet journals. The Lancet. 2018;391(10140):2601. doi:10.1016/S0140-6736(18)31139-5

30. Hornstein P, Tuyishime H, Mutebi M, Lasebikan N, Rubagumya F, Fadelu T. Authorship Equity and Gender Representation in Global Oncology Publications. JCO Glob Oncol. 2022;(8):e2100369. doi:10.1200/GO.21.00369

31. Bendels MHK, Müller R, Brueggmann D, Groneberg DA. Gender disparities in high-quality research revealed by Nature Index journals. Lozano S, ed. PLOS ONE. 2018;13(1):e0189136. doi:10.1371/journal.pone.0189136

32. Wininger AE, Fischer JP, Likine EF, et al. Bibliometric Analysis of Female Authorship Trends and Collaboration Dynamics Over *JBMR*’s 30-Year History. J Bone Miner Res. 2017;32(12):2405–2414. doi:10.1002/jbmr.3232

33. Mamtani M, Shofer F, Mudan A, et al. Quantifying gender disparity in physician authorship among commentary articles in three high-impact medical journals: an observational study. BMJ Open. 2020;10(2):e034056. doi:10.1136/bmjopen-2019-034056

34. Garbern SC, Hyuha G, González Marqués C, et al. Authorship representation in global emergency medicine: a bibliometric analysis from 2016 to 2020. BMJ Glob Health. 2022;7(6):e009538. doi:10.1136/bmjgh-2022-009538

35. Qureshi R, Lê J, Li T, Ibrahim M, Dickersin K. Gender and Editorial Authorship in High-Impact Epidemiology Journals. Am J Epidemiol. 2019;188(12):2140–2145. doi:10.1093/aje/kwz094

36. Misra V, Safi F, Brewerton KA, et al. Gender disparity between authors in leading medical journals during the COVID-19 pandemic: a cross-sectional review. BMJ Open. 2021;11(7):e051224. doi:10.1136/bmjopen-2021-051224

37. Filardo G, Da Graca B, Sass DM, Pollock BD, Smith EB, Martinez MAM. Trends and comparison of female first authorship in high impact medical journals: observational study (1994-2014). BMJ. Published online March 2, 2016:i847. doi:10.1136/bmj.i847

38. Clarivate - data, insights and analytics for the innovation lifecycle. Accessed December 1, 2023. https://clarivate.com/

39. Journal Citations Reports JCR – Clarivate. Accessed December 1, 2023. https://clarivate.com/products/scientific-and-academic-research/research-analytics-evaluation-and-management-solutions/journal-citation-reports/#relatedproducts

40. Publications - Dimensions. Accessed December 1, 2023. https://app.dimensions.ai/discover/publication

41. Telling the gender from a name. Gender Gap in Science. Published July 16, 2018. Accessed March 13, 2024. https://gender-gap-in-science.org/2018/07/16/telling-the-gender-from-a-name/

42. World Bank Country and Lending Groups – World Bank Data Help Desk. Accessed December 1, 2023. https://datahelpdesk.worldbank.org/knowledgebase/articles/906519-world-bank-country-and-lending-groups

43. Sopher CJ, Adamson BJS, Andrasik MP, et al. Enhancing Diversity in the Public Health Research Workforce: The Research and Mentorship Program for Future HIV Vaccine Scientists. Am J Public Health. 2015;105(4):823–830. doi:10.2105/AJPH.2014.302076

44. United Nations Educational, Scientific and Cultural Organization. UNESCO Science Report 2021: The Race Against Time for Smarter Development. United Nations; 2021. doi:10.18356/9789210058575

45. Budden, Amber. Diversity begets diversity: An analysis of relationships between author, reviewer, and editor populations. Eur Sci Ed. 2010;36:31–34.

46. Malekzadeh A, Michels K, Wolfman C, Anand N, Sturke R. Strengthening research capacity in LMICs to address the global NCD burden. Glob Health Action. 13(1):1846904. doi:10.1080/16549716.2020.1846904

47. ESSENCE on Health Research. Health Research Capacity Strengthening in Low and Middle-Income Countries: Current Situation and Opportunities to Leverage Data for Better Coordination and Greater Impact.; 2020.

48. Ghee M, Collins D, Wilson V, Pearson W. The Leadership Alliance: Twenty Years of Developing a Diverse Research Workforce. Peabody J Educ. 2014;89(3):347–367. doi:10.1080/0161956X.2014.913448

49. Munir F, Mason C, McDermott H, Morris J, Bagilhole B, Nevill M. Advancing women’s careers in science, technology, engineering, mathematics and medicine: Evaluating the effectiveness and impact of the Athena SWAN charter. Lond Equal Chall Unit. Published online 2013. Accessed December 1, 2023. https://scholar.google.com/scholar?cluster=4854829323356529127&hl=en&oi=scholarr

50. Ghaffarzadegan N, Hawley J, Desai A. Research Workforce Diversity: The Case of Balancing National versus International Postdocs in US Biomedical Research. Syst Res Behav Sci. 2014;31(2):301–315. doi:10.1002/sres.2190

51. Woolley AW, Chabris CF, Pentland A, Hashmi N, Malone TW. Evidence for a Collective Intelligence Factor in the Performance of Human Groups. Science. 2010;330(6004):686–688. doi:10.1126/science.1193147

52. Launois P, Maher D, Certain E, Ross B, Penkunas MJ. Implementation research training for learners in low- and middle-income countries: evaluating behaviour change after participating in a massive open online course. Health Res Policy Syst. 2021;19(1):59. doi:10.1186/s12961-021-00703-3

