## Supplementary Text, Figures, and Tables for "Does diversity beget diversity? A scientometric analysis of over 150,000 studies and 49,000 authors published in high-impact medical journals between 2007 and 2022"

**Supplemental Text**

**Search query**

Below, you can find the search query that we used to retrieve the raw dataset from DimensionsAI.

*"""search publications where year in [{year}] and journal.id in ["jour.1081531" # JAMA, "jour.1077219" # The Lancet, "jour.1113716" # Nature Medicine, "jour.1034427" # NEJM, "jour.1017377" # The BMJ] return publications[basics+concepts+extras+abstract+categories+authors+authors_count+concepts+concepts_scores+funders+funder_countries+pmid+research_orgs+research_org_names+research_org_countries+research_org_country_names]""".*

**Information on the studied journals**

The five studied journals were: *The Lancet* (created in 1823), *New England Journal of Medicine* (NEJM, created in 1812), *Journal of the American Medical Association* (JAMA, created in 1883), *The British Medical Journal* (BMJ, created in 1840), and *Nature Medicine* (created in 1995).

**Supplemental Tables**

**Table 1. Impact factor (IF) of the considered journals during the study period (2007-2022).**

| **Journal** | **Years with available IF data** | **IF, Mean (SD)** |
| --- | --- | --- |
| NEJM | 2007 – 2021 | 69.5 (32.2) |
| The Lancet | 2007 – 2021 | 55.3 (43.1) |
| JAMA | 2007 – 2021 | 45.5 (32.3) |
| Nature Medicine | 2007 – 2021 | 34 (16.4) |
| The BMJ | 2013 – 2021 | 32.4 (25) |

**Table 2a. Summary statistics about authors, stratified by inferred gender (using an optimistic imputation approach).**

|  | **Inferred gender** | |  |
| --- | --- | --- | --- |
|  | **Female** | **Male** | **P-Value** |
| **N** | 71,212 | 90,920 |  |
| **1+ first-authored paper, N (%)** | 10,129 (14.2) | 14,087 (15.5) | <0.001 |
| **1+ last-authored paper, N (%)** | 10,155 (14.3) | 14,257 (15.7) | <0.001 |
| **2+ first-authored paper, N (%)** | 2,921 (4.1) | 5,418 (6.0) | <0.001 |
| **2+ last-authored paper, N (%)** | 2,893 (4.1) | 5,333 (5.9) | <0.001 |
| **Published in 2+ journals, N (%)** | 10,532 (14.8) | 18,964 (20.9) | <0.001 |
| **Published 2+ papers, N (%)** | 19,816 (27.8) | 31,213 (34.3) | <0.001 |

**Table 2b. Summary statistics about authors, stratified by inferred gender (using a pessimistic imputation approach).**

|  | **Inferred gender** | | |
| --- | --- | --- | --- |
|  | **Female** | **Male** | **P-Value** |
| **N** | 55,364 | 106,768 |  |
| **1+ first-authored paper, N (%)** | 7,850 (14.2) | 16,366 (15.3) | <0.001 |
| **1+ last-authored paper, N (%)** | 7,936 (14.3) | 16,476 (15.4) | <0.001 |
| **2+ first-authored paper, N (%)** | 2,405 (4.3) | 5,934 (5.6) | <0.001 |
| **2+ last-authored paper, N (%)** | 2,372 (4.3) | 5,854 (5.5) | <0.001 |
| **Published in 2+ journals, N (%)** | 8,677 (15.7) | 20,819 (19.5) | <0.001 |
| **Published 2+ papers, N (%)** | 15,948 (28.8) | 35,081 (32.9) | <0.001 |

**Table 3a. Summary statistics about authors, stratified by country income category (using an optimistic imputation approach).**

|  | **Country income category** | |  |
| --- | --- | --- | --- |
|  | **HIC** | **LMIC** | **P-Value** |
| **N** | 140,674 | 21,458 |  |
| **1+ first-authored paper, N (%)** | 21,142 (15.0) | 3,074 (14.3) | 0.007 |
| **1+ last-authored paper, N (%)** | 21,245 (15.1) | 3,167 (14.8) | 0.194 |
| **2+ first-authored paper, N (%)** | 7,605 (5.4) | 734 (3.4) | <0.001 |
| **2+ last-authored paper, N (%)** | 7,513 (5.3) | 713 (3.3) | <0.001 |
| **Published in 2+ journals, N (%)** | 26,690 (19.0) | 2,806 (13.1) | <0.001 |
| **Published 2+ papers, N (%)** | 45,638 (32.4) | 5,391 (25.1) | <0.001 |

*LMIC: low- or middle-income country, HIC: high-income country.

**Table 3b. Summary statistics about authors, stratified by country income category (using a pessimistic imputation approach).**

|  | **Country income category** | |  |
| --- | --- | --- | --- |
|  | **HIC** | **LMIC** | **P-Value** |
| **N** | 143,298 | 18,834 |  |
| **1+ first-authored paper, N (%)** | 21,583 (15.1) | 2,633 (14.0) | <0.001 |
| **1+ last-authored paper, N (%)** | 21,704 (15.1) | 2,708 (14.4) | 0.006 |
| **2+ first-authored paper, N (%)** | 7,622 (5.3) | 717 (3.8) | <0.001 |
| **2+ last-authored paper, N (%)** | 7,540 (5.3) | 686 (3.6) | <0.001 |
| **Published in 2+ journals, N (%)** | 26,704 (18.6) | 2,792 (14.8) | <0.001 |
| **Published 2+ papers, N (%)** | 45,785 (32.0) | 5,244 (27.8) | <0.001 |

*LMIC: low- or middle-income country, HIC: high-income country.

**Table 4a. Representation of female authors in publications, when filtering for articles with a maximum of 30 authors, across three missing data imputation scenarios (normal, optimistic, and pessimistic approaches).**

| **Journal \ Scenario** | **Normal (%)** | **Optimistic (%)** | **Pessimistic (%)** |
| --- | --- | --- | --- |
| NEJM | 31.0 | 36.4 | 28.6 |
| The Lancet | 32.2 | 37.9 | 29.5 |
| **Overall** | 33.1 | 38.2 | 30.5 |
| JAMA | 33.9 | 38.5 | 31.6 |
| Nature Medicine | 34.0 | 39.8 | 31.0 |
| The BMJ | 34.9 | 39.0 | 32.8 |

**Table 4b. Representation of female authors in publications, when filtering for articles with a maximum of 40 authors, across three missing data imputation scenarios (normal, optimistic, and pessimistic approaches).**

| **Journal \ Scenario** | **Normal (%)** | **Optimistic (%)** | **Pessimistic (%)** |
| --- | --- | --- | --- |
| NEJM | 31.1 | 36.6 | 28.6 |
| The Lancet | 32.2 | 37.9 | 29.5 |
| **Overall** | 33.2 | 38.4 | 30.6 |
| JAMA | 33.9 | 38.5 | 31.5 |
| Nature Medicine | 34.3 | 40.1 | 31.3 |
| The BMJ | 35.0 | 39.1 | 32.8 |

**Table 4c. Representation of female author in publications, when filtering for articles with a maximum of 50 authors, across three missing data imputation scenarios (normal, optimistic, and pessimistic approaches).**

| **Journal \ Scenario** | **Normal (%)** | **Optimistic (%)** | **Pessimistic (%)** |
| --- | --- | --- | --- |
| NEJM | 31.1 | 36.6 | 28.6 |
| The Lancet | 32.2 | 37.9 | 29.5 |
| **Overall** | 33.2 | 38.4 | 30.6 |
| JAMA | 33.9 | 38.5 | 31.5 |
| Nature Medicine | 34.3 | 40.1 | 31.3 |
| The BMJ | 35.0 | 39.1 | 32.8 |

**Table 5. Summary statistics about team composition, with no conditioning (baseline) and with conditioning to assess whether diversity begets diversity.**

|  | **p (A \| B)** | **B** | | | | | | |
| --- | --- | --- | --- | --- | --- | --- | --- | --- |
|  | **N (%)** | **If first author is female** | **If last author is female** | **If first author is LMIC** | **If last author is LMIC** | **If at least 1 female author** | **If at least 1 LMIC author** | **Baseline (overall)** |
| **A** | **First author is female** | **—** | **6,184 (40.1)** | 1,371 (31.5) | 1,317 (30.4) | **15,566 (47.0)** | 2,469 (32.3) | 15,566 (31.3) |
|  | **Last author is female** | **6,184 (39.7)** | **—** | 1,407 (32.3) | 1,405 (32.4) | **15,436 (46.6)** | 2,433 (31.9) | 15,566 (31.3) |
|  | **First author is LMIC** | 1,371 (8.8) | 1,407 (9.1) | — | **3.057 (70.5)** | 2,952 (8.9) | **4,359 (57.1)** | 4,359 (8.8) |
|  | **Last author is LMIC** | 1,317 (8.5) | 1,405 (9.1) | **3,057 (70.1)** | — | 2,919 (8.8) | **4,337 (56.8)** | 4,337 (8.7) |
|  | **At least 1 female author** | 15,566 (100) | 15,436 (100) | 2,952 (67.7) | 2,919 (67.3) | — | **5,661 (74.2)** | 33,129  (66.6) |
|  | **At least 1 LMIC author** | 2,469 (15.9) | 2,433 (15.8) | 4,359 (100) | 4,337 (100) | **5,661 (17.1)** | — | 7,633 (15.3) |
|  | **At least 2 female authors** | **9,915 (63.7)** | **9,800 (63.5)** | **1,749 (40.1)** | 1,681 (38.8) | **18,687 (56.4)** | **3,767 (49.4)** | 18,687 (37.6) |
|  | **At least 2 LMIC authors** | 1,646 (10.6) | 1,689 (10.9) | **3,728 (85.5)** | **3,707 (85.5)** | **3,870 (11.7)** | **5,172 (67.8)** | 5,172 (10.4) |

***** Each cell represents a conditional probability p(A|B) (e.g., the first row corresponds to the conditional probability p(First author is female | First author is female)), expressed as a percentage value. Statistically significant values are represented in bold and refer to a p-value < 0.002 after performing a Chi-squared test (e.g., *the first author is female* vs. *the first author is male*). For simplicity, we report statistics for one category only (female for inferred gender and LMIC for country income). A Bonferroni correction was applied considering 20 tests and a familywise error rate of 0.05.
White cells correspond to cases without sufficient evidence to reject the null hypothesis; **green cells** correspond to statistically significant relationships; **gray cells** correspond to statistically significant relationships between related events (e.g., the event “first author being female” is not independent from the event “having at least one female author”) and are thus not as relevant.

**Table 6a. Baseline probabilities, stratified by team size.**

| **Team Size** | **Fraction of authors** | **P (Female author)** | **P (LMIC author)** |
| --- | --- | --- | --- |
| 2-3 authors | 0.234 | 0.288 | 0.075 |
| 4-5 authors | 0.122 | 0.335 | 0.107 |
| 6+ authors | 0.644 | 0.346 | 0.102 |

* LMIC:	low- or middle-income country.
* Information about missing data can be found in Table 2.

**Table 6b. Baseline probabilities, stratified by time period.**

| **Time period** | **Fraction of authors** | **P (Female author)** | **P (LMIC author)** |
| --- | --- | --- | --- |
| Before December 2016, inclusive | 0.442 | 0.304 | 0.083 |
| Between January 2017 and December 2019, inclusive | 0.315 | 0.337 | 0.106 |
| January 2020 onwards | 0.243 | 0.372 | 0.107 |

**Table 7. Sensitivity analysis of Figure 3, restricted to publications whose authors’ gender could be inferred, stratified by team size and by time period.**

| **X** | p ( first author \| **X** ) | p ( last author \| **X** ) |
| --- | --- | --- |
| **Team Size** | **Any** | |
| Female | 0.15 (0.147, 0.154) | 0.151 (0.147, 0.151) |
| Male | 0.13 (0.128, 0.133) | 0.13 (0.128, 0.13) |
| LMIC | 0.12 (0.115, 0.124) | 0.119 (0.115, 0.119) |
| HIC | 0.131 (0.13, 0.132) | 0.131 (0.13, 0.131) |
| **Team Size** | **3+ authors** | |
| Female | 0.13 (0.127, 0.134) | 0.131 (0.127, 0.131) |
| Male | 0.118 (0.116, 0.12) | 0.118 (0.116, 0.118) |
| LMIC | 0.11 (0.105, 0.114) | 0.11 (0.105, 0.11) |
| HIC | 0.115 (0.115, 0.116) | 0.115 (0.115, 0.115) |
| **Team Size** | **2 - 3 authors** | |
| Female | 0.385 (0.374, 0.396) | 0.385 (0.375, 0.385) |
| Male | 0.387 (0.382, 0.392) | 0.387 (0.382, 0.387) |
| LMIC | 0.36 (0.325, 0.393) | 0.36 (0.328, 0.36) |
| HIC | 0.362 (0.358, 0.367) | 0.362 (0.358, 0.362) |
| **Team Size** | **4 - 5 authors** | |
| Female | 0.237 (0.225, 0.248) | 0.237 (0.226, 0.237) |
| Male | 0.237 (0.23, 0.243) | 0.237 (0.231, 0.237) |
| LMIC | 0.221 (0.198, 0.243) | 0.222 (0.199, 0.222) |
| HIC | 0.223 (0.221, 0.226) | 0.223 (0.221, 0.223) |
| **Team Size** | **6+ authors** | |
| Female | 0.088 (0.085, 0.091) | 0.088 (0.085, 0.088) |
| Male | 0.087 (0, 0.089) | 0.087 (0.085, 0.087) |
| LMIC | 0.08 (0.075, 0.085) | 0.08 (0.076, 0.08) |
| HIC | 0.079 (0.079, 0.08) | 0.079 (0.079, 0.079) |
| **Time Period** | **Before December 2016 (inclusive)** | |
| Female | 0.178 (0.172, 0.183) | 0.178 (0.172, 0.178) |
| Male | 0.193 (0.19, 0.196) | 0.192 (0.19, 0.192) |
| LMIC | 0.124 (0.115, 0.132) | 0.124 (0.116, 0.124) |
| HIC | 0.142 (0.14, 0.143) | 0.142 (0.14, 0.142) |
| **Time Period** | **January 2017 - December 2019 (inclusive)** | |
| Female | 0.165 (0.159, 0.172) | 0.165 (0.158, 0.165) |
| Male | 0.176 (0.173, 0.18) | 0.176 (0.172, 0.176) |
| LMIC | 0.123 (0.115, 0.133) | 0.123 (0.113, 0.123) |
| HIC | 0.125 (0.124, 0.127) | 0.125 (0.124, 0.125) |
| **Time Period** | **January 2020 onwards** | |
| Female | 0.157 (0.15, 0.162) | 0.157 (0.151, 0.157) |
| Male | 0.163 (0.158, 0.166) | 0.163 (0.159, 0.163) |
| LMIC | 0.109 (0.1, 0.118) | 0.11 (0.101, 0.11) |
| HIC | 0.119 (0.118, 0.121) | 0.119 (0.118, 0.119) |

* LMIC: low- or middle-income country, HIC: high-income country.

**Table 8. Sensitivity analysis of Figure 3, restricted to publications whose authors’ affiliation and country income category was available, stratified by team size and by time period.**

| **X** | p ( first author \| **X** ) | p ( last author \| **X** ) |
| --- | --- | --- |
| **Team Size** | **Any** | |
| Female | 0.116 (0.108, 0.124) | 0.115 (0.108, 0.115) |
| Male | 0.117 (0.114, 0.121) | 0.118 (0.114, 0.118) |
| LMIC | 0.118 (0.112, 0.125) | 0.119 (0.112, 0.119) |
| HIC | 0.100 (0.096, 0.103) | 0.099 (0.096, 0.099) |
| **Team Size** | **3+ authors** | |
| Female | 0.108 (0.100, 0.115) | 0.108 (0.101, 0.108) |
| Male | 0.109 (0.105, 0.113) | 0.109 (0.106, 0.109) |
| LMIC | 0.108 (0.101, 0.116) | 0.108 (0.1, 0.108) |
| HIC | 0.093 (0.089, 0.097) | 0.093 (0.089, 0.093) |
| **Team Size** | **2 - 3 authors** | |
| Female | 0.379 (0.33, 0.425) | 0.379 (0.333, 0.379) |
| Male | 0.381 (0.354, 0.406) | 0.381 (0.355, 0.381) |
| LMIC | 0.364 (0.331, 0.399) | 0.362 (0.327, 0.362) |
| HIC | 0.357 (0.329, 0.384) | 0.359 (0.332, 0.359) |
| **Team Size** | **4 - 5 authors** | |
| Female | 0.243 (0.214, 0.272) | 0.244 (0.214, 0.244) |
| Male | 0.240 (0.227, 0.256) | 0.240 (0.225, 0.24) |
| LMIC | 0.220 (0.194, 0.249) | 0.221 (0.192, 0.221) |
| HIC | 0.220 (0.201, 0.241) | 0.220 (0.199, 0.22) |
| **Team Size** | **6+ authors** | |
| Female | 0.084 (0.077, 0.09) | 0.083 (0.077, 0.083) |
| Male | 0.083 (0.080, 0.086) | 0.083 (0.080, 0.083) |
| LMIC | 0.079 (0.073, 0.084) | 0.079 (0.074, 0.079) |
| HIC | 0.073 (0.069, 0.075) | 0.073 (0.07, 0.073) |
| **Time Period** | **Before December 2016 (inclusive)** | |
| Female | 0.124 (0.112, 0.137) | 0.124 (0.111, 0.124) |
| Male | 0.130 (0.125, 0.137) | 0.130 (0.124, 0.13) |
| LMIC | 0.129 (0.118, 0.139) | 0.129 (0.119, 0.129) |
| HIC | 0.111 (0.105, 0.117) | 0.112 (0.106, 0.112) |
| **Time Period** | **January 2017 - December 2019 (inclusive)** | |
| Female | 0.111 (0.098, 0.124) | 0.111 (0.098, 0.111) |
| Male | 0.112 (0.105, 0.118) | 0.112 (0.105, 0.112) |
| LMIC | 0.114 (0.103, 0.124) | 0.114 (0.104, 0.114) |
| HIC | 0.093 (0.087, 0.099) | 0.093 (0.087, 0.093) |
| **Time Period** | **January 2020 onwards** | |
| Female | 0.110 (0.098, 0.121) | 0.110 (0.099, 0.11) |
| Male | 0.105 (0.097, 0.112) | 0.105 (0.097, 0.105) |
| LMIC | 0.109 (0.098, 0.121) | 0.109 (0.098, 0.109) |
| HIC | 0.091 (0.085, 0.097) | 0.091 (0.084, 0.091) |

* LMIC: low- or middle-income country, HIC: high-income country.

**Supplemental Figures**

**Figure 1. Time series representing the evolution of each considered journal’s impact factor over the study period.**

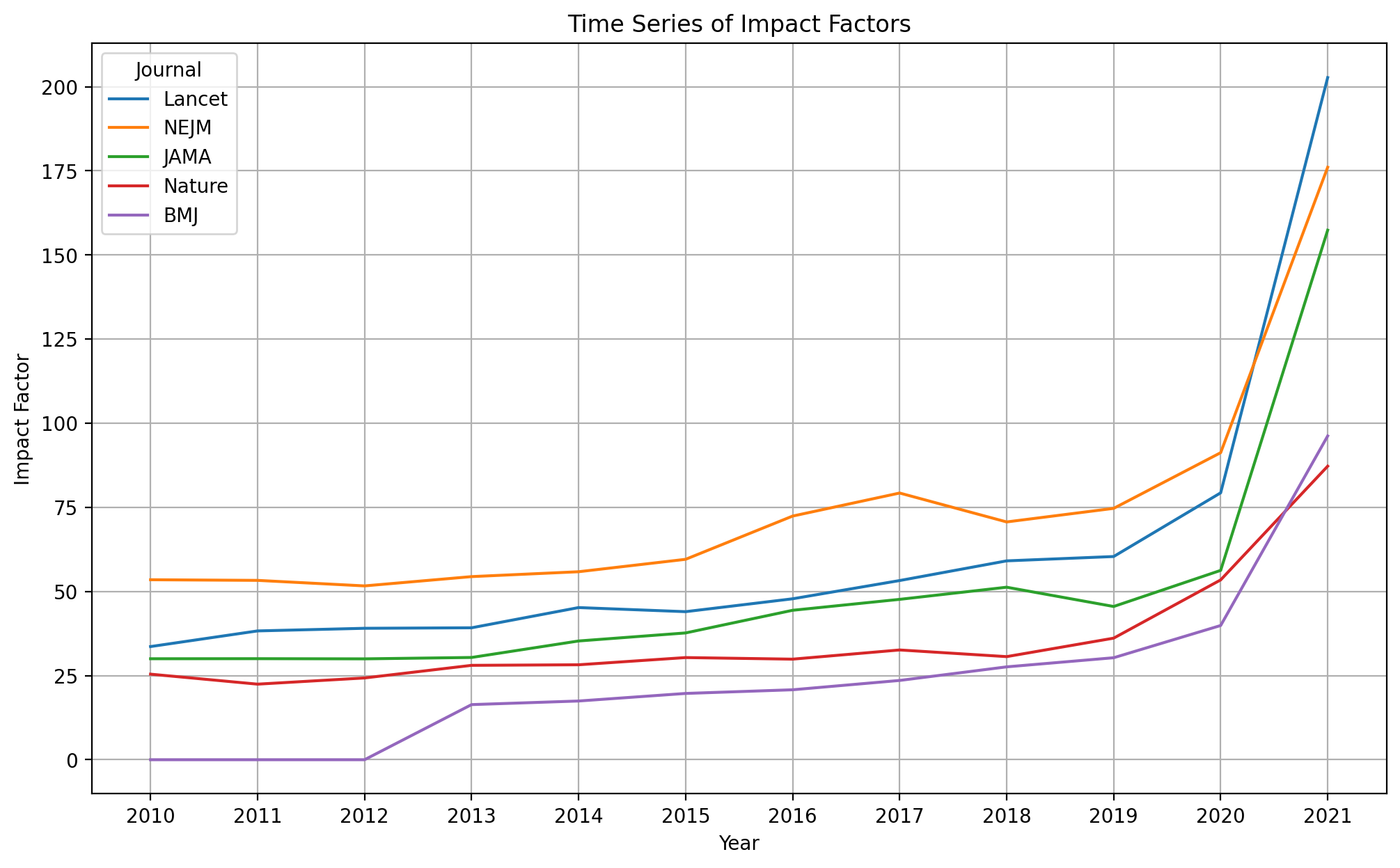

**Figure 2a. Proportion of authors by continent.**

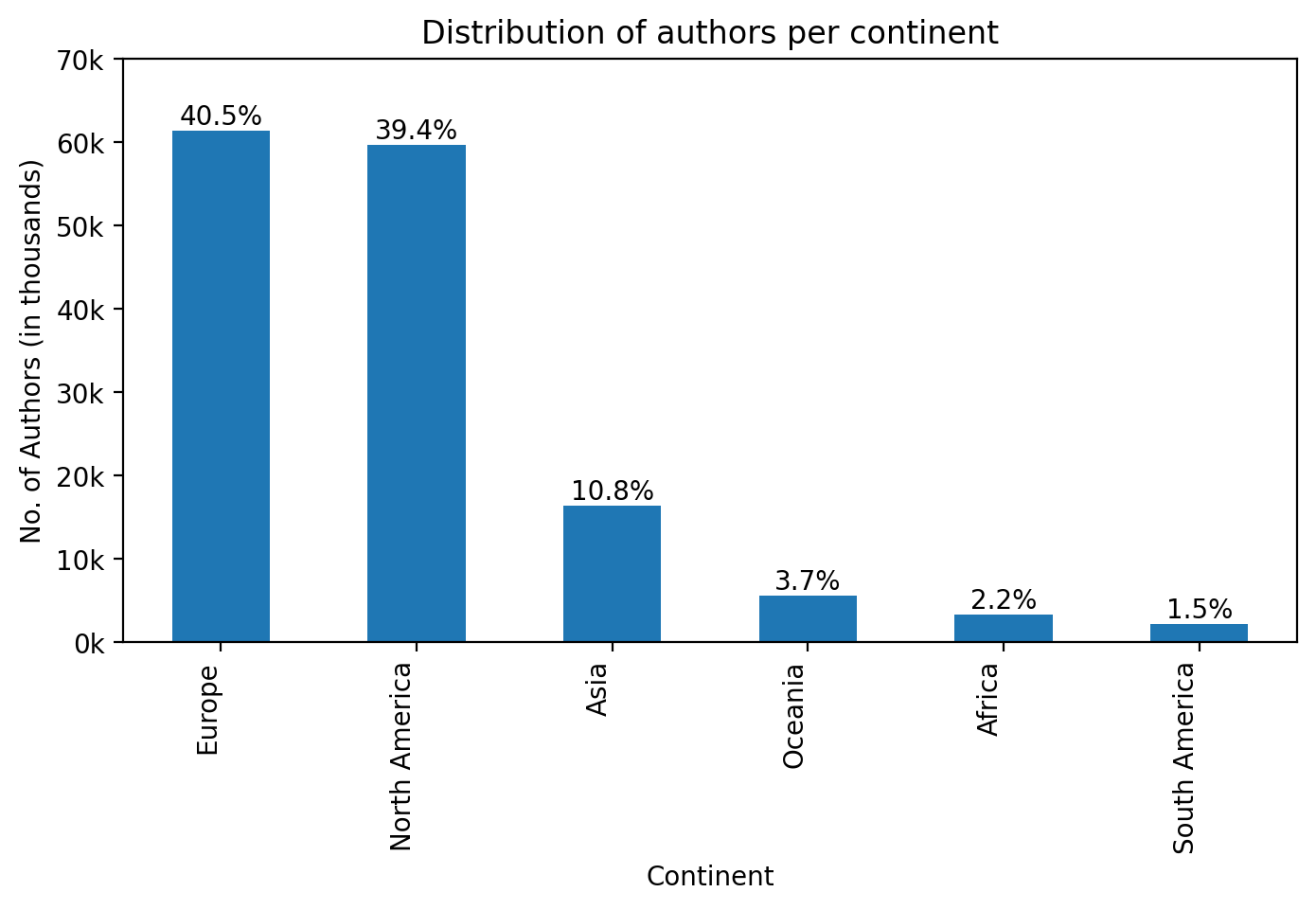

**Figure 2b. Distribution of the number of authors by country (top 20).**

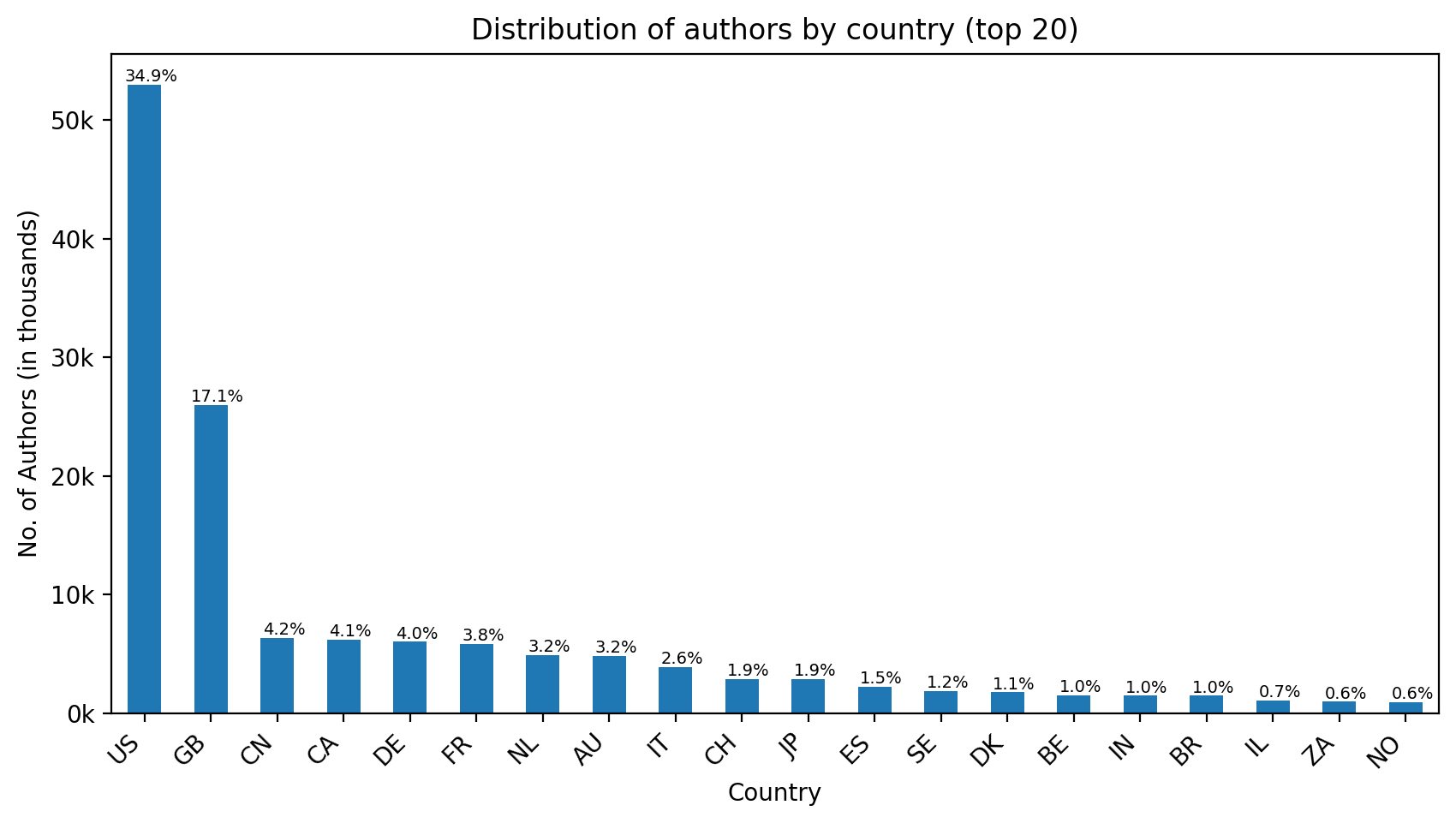

**Figure 3a. Geographical diversity, stratified by journal, when using an optimistic missing data imputation approach (i.e., every author with missing geographical information is considered as affiliated with an institution located in a low- or middle-income country).**

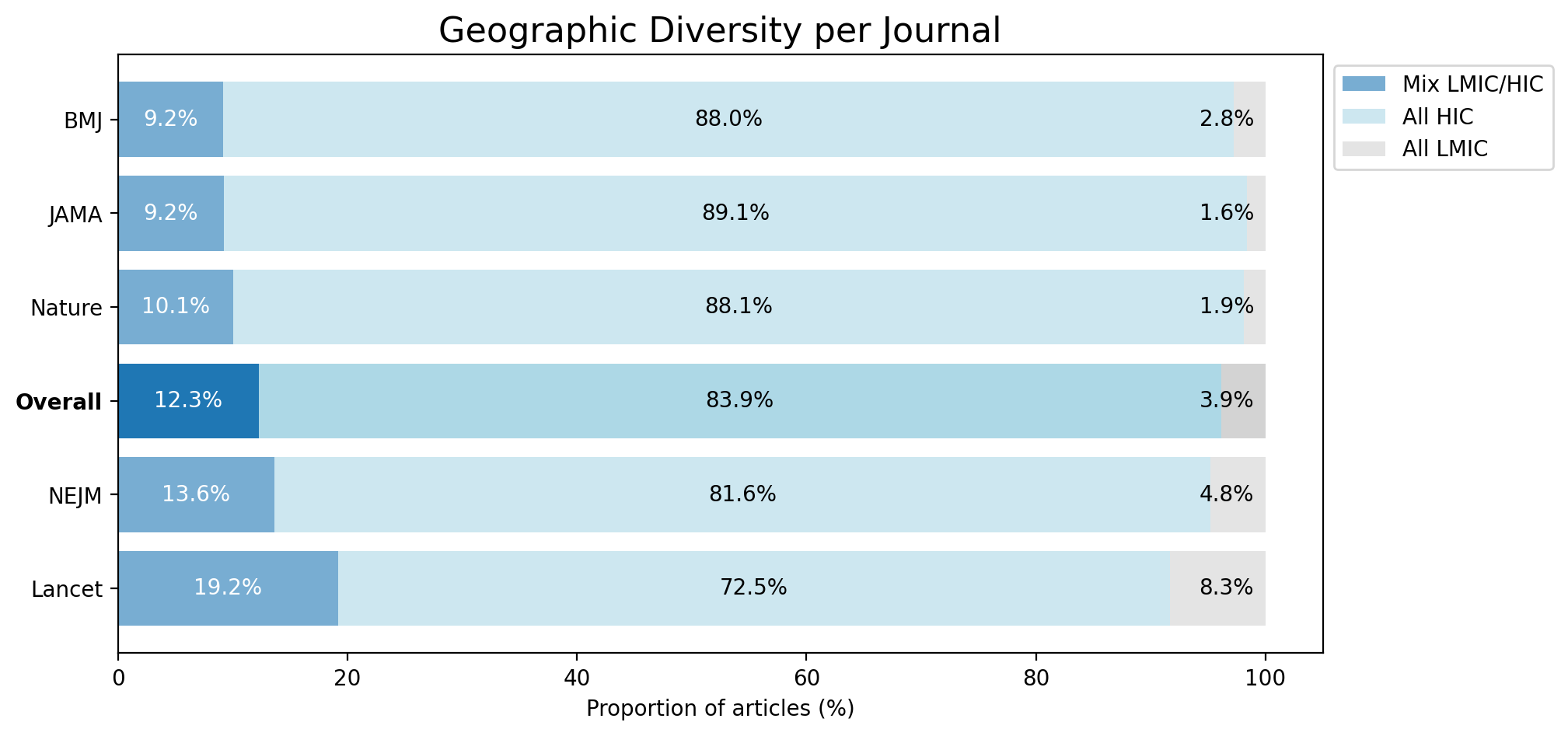

**Figure 3b. Geographical diversity, stratified by journal, when using a pessimistic missing data imputation approach (i.e., every author with missing geographical information is considered as affiliated with an institution located in a high-income country).**

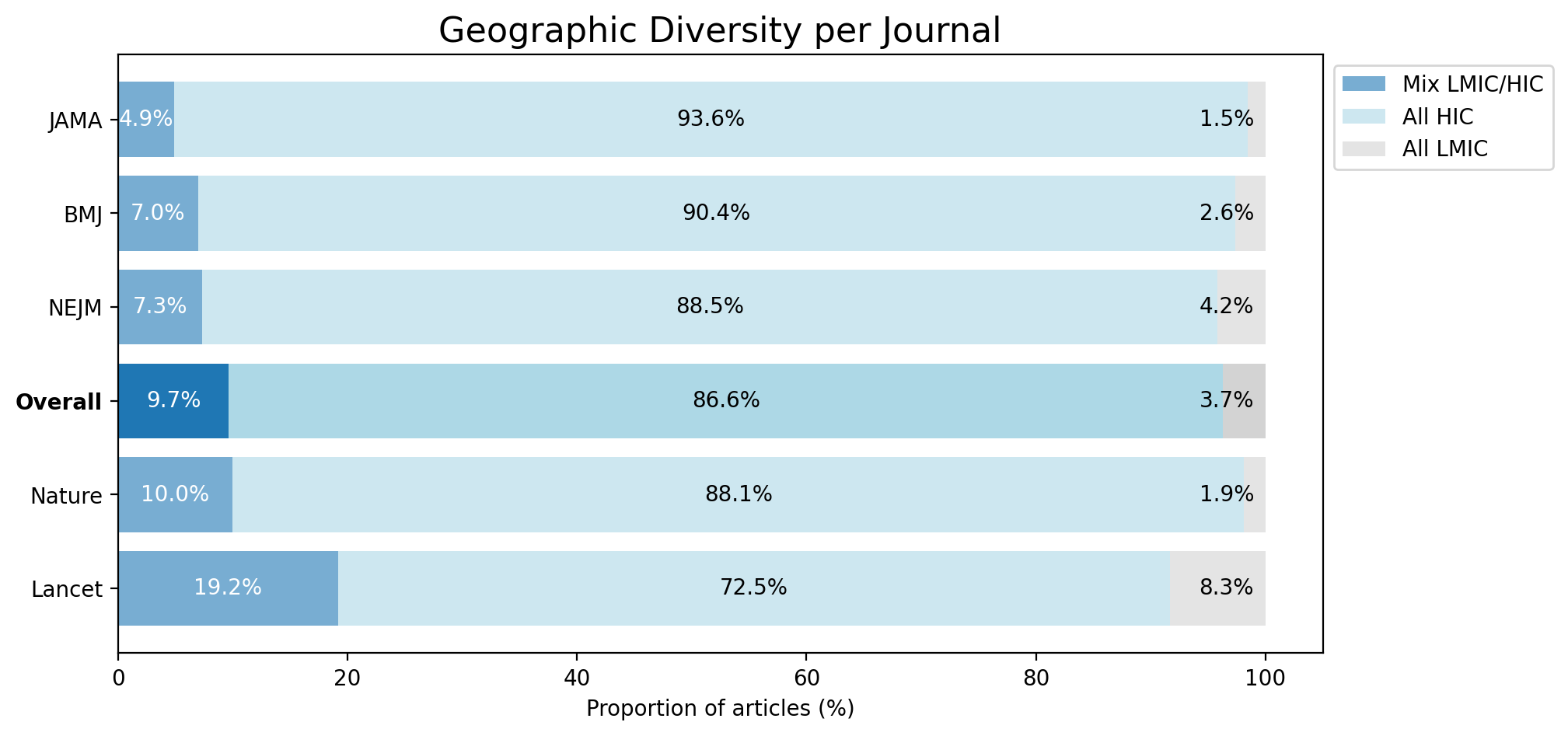
